# Residual SARS-CoV-2 viral antigens detected in gastrointestinal and hepatic tissues from two recovered COVID-19 patients

**DOI:** 10.1101/2020.10.28.20219014

**Authors:** Chun Chau Lawrence Cheung, Denise Goh, Xinru Lim, Tracy Zhijun Tien, Jeffrey Chun Tatt Lim, Sanjna Nilesh Nerurkar, Loong Shihleone, Peng Chung Cheow, Chung Yip Chan, Ye Xin Koh, Thuan Tong Tan, Shirin Kalimuddin, Wai Meng David Tai, Jia Lin Ng, Jenny Guek Hong Low, Joe Yeong, Tony Kiat Hon Lim

## Abstract

Residual SARS-CoV-2 RNA has been detected in stool samples and gastrointestinal tissues during the convalescence phase of COVID-19 infection. This raises concern for persistence of SARS-CoV-2 virus particles and faecal-oral transmissibility in recovered COVID-19 patients. Using multiplex immunohistochemistry, we unexpectedly detected SARS-CoV-2 viral antigens in intestinal and liver tissues, in surgical samples obtained from two patients who recovered from COVID-19. We further validated the presence of virus by RT-PCR and flow cytometry to detect SARS-CoV-2-specific immunity in the tissues. These findings might have important implications in terms of disease management and public health policy regarding transmission of COVID-19 via faecal-oral and iatrogenic routes during the convalescence phase.

## Introduction

Coronavirus disease 2019 (COVID-19), caused by the novel coronavirus SARS-CoV-2, was first reported in December 2019 and has quickly become a global pandemic. Direct SARS-CoV-2 viral infection of gastrointestinal organs and consequential faecal-oral transmission route is a concern in COVID-19 pathology. SARS-CoV-2 RNA has been detected in stool samples during active and convalescence phase of infection (1, 2), suggesting that faecal-oral viral shedding might occur during COVID-19 convalescence (3). However, no report has yet detected viral antigens within gastrointestinal and hepatic organs during the convalescent phase.

## Materials and Methods

### Study cohort

Patient 1 was a middle-aged male who was diagnosed with COVID-19 in April 2020. He experienced mild upper respiratory tract symptoms, and was confirmed COVID-19 negative by two consecutive nasopharyngeal swabs in May 2020. During the hospitalisation, he was incidentally found to have a large circumferential malignant mass in the colon. He underwent laparoscopic right hemicolectomy 9 days after testing negative for COVID-19.

Patient 2 was a middle-aged male who was diagnosed with COVID-19 pneumonia in June 2020. Prior to the diagnosis, he had a fever with no attending respiratory symptoms. He had a significant medical history of chronic hepatitis B virus-related liver cirrhosis and segment VII hepatocellular carcinoma (HCC). The patient was confirmed COVID-19 negative by two consecutive nasopharyngeal swabs in mid-June 2020. He underwent curative resection of HCC in August 2020, 85 days after being testing negative for COVID-19.

This study was approved by the SingHealth Centralised Institutional Review Board (reference number: 2019/2653). Written informed consent was obtained from both patients 1 and 2.

### Specimen collection

Colon, ileum, appendix and lymph nodes tissues were removed during surgery from patient 1, while liver tissues were removed from patient 2. The tissues were formalin-fixed paraffin-embedded (FFPE) for analysis.

### Immunohistochemistry

Immunohistochemistry was performed on FFPE tissue samples as previously described.(4-6) In brief, FFPE tissue sections (4-µm thick) were labelled with antibodies targeting the SARS-CoV-2 nucleocapsid (Polyclonal) and spike protein (1A9). Appropriate positive and negative controls were included. Nuclei were counterstained with haematoxylin. Images were captured using an IntelliSite Ultra-Fast Scanner (Philips, Netherlands).

### Multiplex immunohistochemistr

Multiplex immunohistochemistry was performed using an Opal Multiplex fIHC kit (Akoya Bioscience, USA), as we previously described.(7-9) In brief, FFPE tissue samples (4 µm-thick) were labelled with primary antibodies against the SARS-CoV-2 nucleocapsid (Polyclonal), ACE2 (EPR4435(2)), CD14 (EPR3653) and CD68 (PG-M1) (Table 1), followed by appropriate secondary antibodies and Opal fluorophore-conjugated tyramide signal amplification (Akoya Bioscience, USA). Finally, the samples were incubated with the nuclear counterstain spectral DAPI (Akoya Bioscience, USA). Images were captured under a Vectra 3 pathology imaging system microscope (PerkinElmer, Inc., USA) and analysed using inForm (Akoya Bioscience, USA) and HALO™ (Indica Labs) software.

**Table 1.** Details of antibodies used for immunohistochemistry, multiplex immunohistochemistry and flow cytometry.

| **Primary antibody** | **Company** | **Catalog #** | **Host species** | **Clone** |
| --- | --- | --- | --- | --- |
| SARS-CoV-2 (NP) | Novus Biologicals | NB100-56576 | Rabbit | Polyclonal |
| Spike protein | GeneTex | GTX632604 | Mouse | 1A9 |
| ACE2 | Abcam | Ab108252 | Rabbit | EPR4435(2) |
| CD14 | Abcam | ab133335 | Rabbit | EPR3653 |
| CD68 | Dako | DKO.M087601 | Mouse | PG-M1 |
| CD45 | Invitrogen | MHCD4530 | Mouse | HI30 |
| CD103 | BD Biosciences | 743652 | Mouse | Ber-ACT8 |
| CD4 | BD Biosciences | 566355 | Mouse | SK3 |
| CD3 | Invitrogen | 58-0038-42 | Mouse | UCHT1 |
| CD39 | BioLegend | 563678 | Mouse | TU66 |
| Granzyme B | Invitrogen | GRB18 | Mouse | GB11 |
| CD38 | Invitrogen | 46-0388-42 | Mouse | HB7 |

### RNA extraction and RT-PCR

Total RNA was extracted using the AllPrep FFPE DNA/RNA Kit (Qiagen, Germany) according to the manufacturer’s protocol. Extracted RNA was tested for SARS-CoV-2 using the RealStar® SARS-Cov-2 RT-PCR kit 1.0 (Altona Diagnostics, Germany) following the manufacturer’s instructions.

### Ex vivo peptide stimulation assay

To simulate SARS-CoV-2 infection and to examine SARS-CoV-2-specific immune cells, SARS-CoV-2 PepTivator peptide pools (Miltenyi Biotec, Germany) were used. Lyophilised peptide pools were reconstituted as per manufacturer’s instruction. Blood and tissue samples were prepared as previously described.(8) 1×10^6^ cells were stimulated with 1 μg/mL peptides for 16 hours at 37°C in 5% CO_2_ in RPMI 1640 media (Gibco, USA) supplemented with 10% FBS (Hyclone) and 1% Penicillin-Streptomycin-Glutamine (Gibco, USA).(10, 11) Negative controls were left unstimulated. Brefeldin A (1 μg/ml, Sigma Aldrich, Germany) was added 2 hours into the stimulation assay.

### Flow cytometry and Analysis

Cells were stained with Zombie NIR Fixable Viability dye (BioLegend, USA) for 10 min at 4°C in the dark, washed with PBS at 300 g for 5 min at 4°C, and blocked with Human TruStain FcX (BioLegend, USA) for 10 min at room temperature prior to staining with fluorescent-conjugated antibodies. Next, cells were surface stained with antibody cocktail containing Pacific Orange-anti CD45 (HI30), BV605-anti CD103 (Ber-ACT8), BV750-anti CD4 (SK3), Alexa 532-anti CD3 (UCHT1), PerCP-eFluor 710-anti CD38 (HB7) and PE-CF594-anti CD39 (TU66) (Table 1) in Brilliant Stain Buffer (BD Biosciences, USA), followed by incubation for 30 min at 4°C in the dark.

Prior to intracellular staining, cells were fixed and permeabilised with BD Cytofix/Cytoperm™ (BD Biosciences, USA). Cells were then stained with PE-Cy5.5-anti Granzyme B (GB11) (Table 1) in Brilliant Stain Buffer (BD Biosciences, USA), for 30 min at 4°C in the dark. Finally, cells were washed with 1x BD Perm/Wash Buffer™ (BD Biosciences, USA) and resuspended in PBS containing 2% FBS for flow cytometry using Cytek Aurora spectral flow cytometer (Cytek Biosciences, USA). Data analysis was performed using FlowJo V.10 software (FlowJo LLC, USA).

## Results

Using conventional immunohistochemistry, we detected the SARS-CoV-2 nucleocapsid in the intestinal and liver tissues from patients 1 and 2, respectively. For patient 1, we observed positive staining for the viral antigen in adjacent normal colonic crypts and polyps (Figure 1A, B), as well as lymph nodes, ileum, and appendix (Figure 2A-C), but not in the intratumour region (Figure 1C). We confirmed the presence of viral RNA by RT-PCR. We also observed positive staining for the viral antigen in some of the scattered regions of benign hepatocytes and HCC cells particularly at the invasive margin of the liver sample taken from patient 2 (Figure 1D-F). More importantly, immune cells, including sinusoidal Kupffer cells, were positively stained for the viral antigen (Figure 1D). Of note, we were unable to detect viral RNA in liver tissues, possibly because almost 3 months had passed since recovery. Furthermore, multiplex immunohistochemistry staining showed that the SARS-CoV-2 nucleocapsid and angiotensin converting enzyme 2 (ACE2) receptor were in close proximity in both colon and liver cells (Figure 1G, I). We also discovered that some viral antigen-positive cells were positive for CD14 and CD68 (Figure 1H, J), suggesting that these were likely monocytes and macrophages. Validating our findings, we detected the SAR-CoV-2 spike protein in the abovementioned tissues from both patients (Figure 2D-H). Lastly, to examine SARS-CoV-2-specific immunity in the tissues from recovered patients, we performed *ex vivo* peptide stimulation assays involving a cocktail of the viral nucleocapsid, spike and membrane proteins. Notably, SARS-CoV-2-specific CD38^+^Granzyme B^+^CD4^+^ T cells were elicited from the tissues in a comparable fashion with matched blood samples (Figure 2I, J), suggesting that SARS-CoV-2-specific memory T cells may be maintained in both tissue and blood.

**Figure 1.**
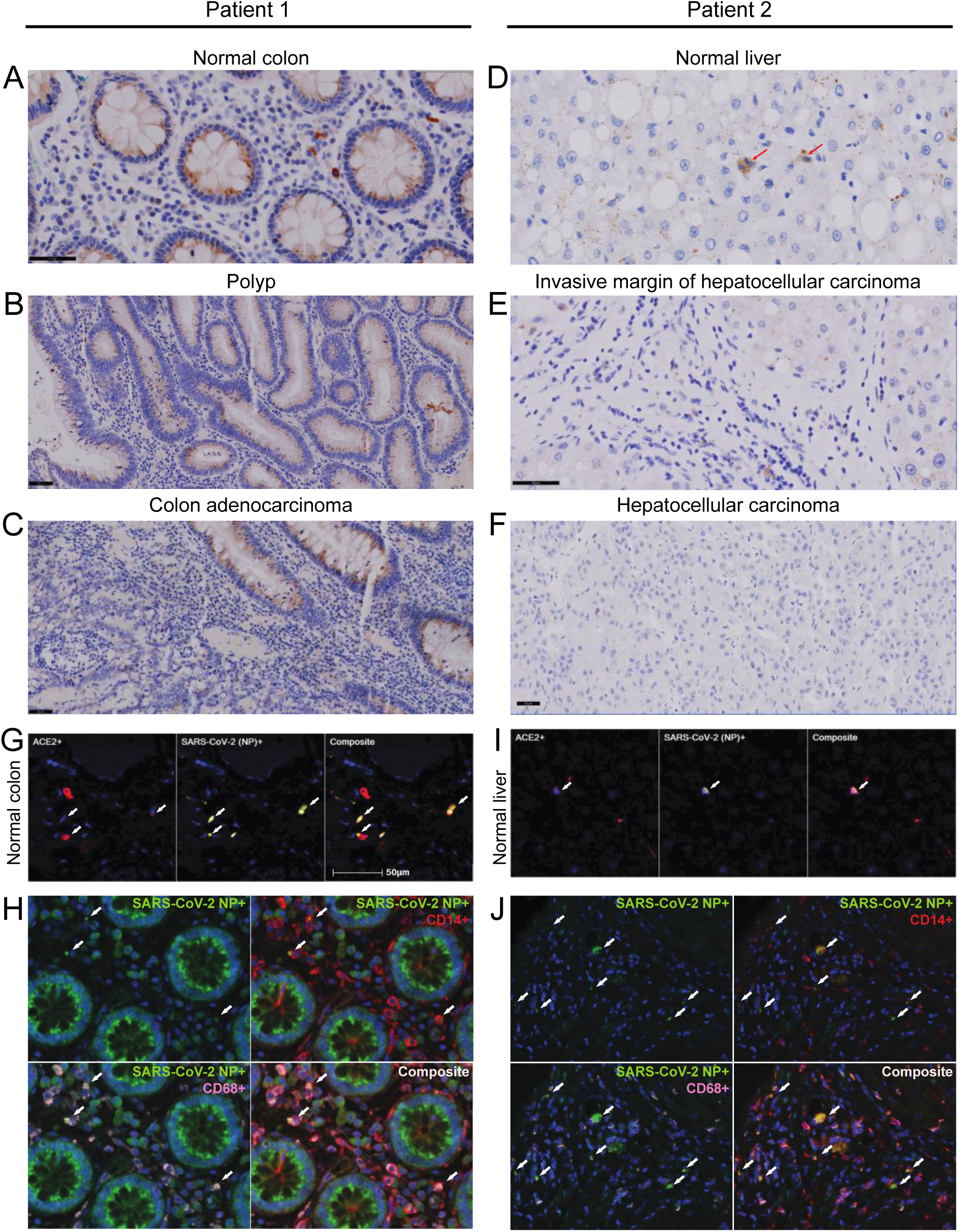
Immunohistochemical staining of the SARS-CoV-2 nucleocapsid protein in normal colon and liver tissue. (A-B) Positive SARS-CoV-2 nucleocapsid staining in colonic crypts (A) and polyps (B), both with a granular supranuclear cytoplasmic pattern. (C) Positive SARS-CoV-2 nucleocapsid staining in normal colonic crypts (top right) but negative staining in the adjacent neoplastic glands (bottom left). (D) Positive SARS-CoV-2 nucleocapsid staining in benign hepatocytes and sinusoidal Kupffer cells (red arrows). (E) Numerous SARS-CoV-2 viral antigen-positive cells were seen along the invasive margin of HCC, including the tumour and immune cells. (F) Negative SARS-CoV-2 nucleocapsid staining in the centre of HCC. (G) Multiplex immunohistochemistry of normal colon tissue. From left to right: ACE2 (red), SARS-CoV-2 nucleocapsid (yellow), and composite. The white arrows indicate co-localisation. Magnification x200. (H) Multiplex immunohistochemistry of normal colon tissue. From left to right: SARS-CoV-2 nucleocapsid (green), SARS-CoV-2 nucleocapsid (green) with CD14 (red), SARS-CoV-2 nucleocapsid (green) with CD68 (pink), and composite. The white arrows indicate co-localisation. (I) Multiplex immunohistochemistry of normal liver tissue. From left to right: ACE2 (red), SARS-CoV-2 nucleocapsid (yellow), and composite. The white arrows indicate co-localisation. Magnification x200. (J) Multiplex immunohistochemistry of normal liver tissue. From left to right: SARS-CoV-2 nucleocapsid (green), SARS-CoV-2 nucleocapsid (green) with CD14 (red), SARS-CoV-2 nucleocapsid (green) with CD68 (pink), and composite. The white arrows indicate co-localisation. Scale bar, 50 μm (A-J).

**Figure 2.**
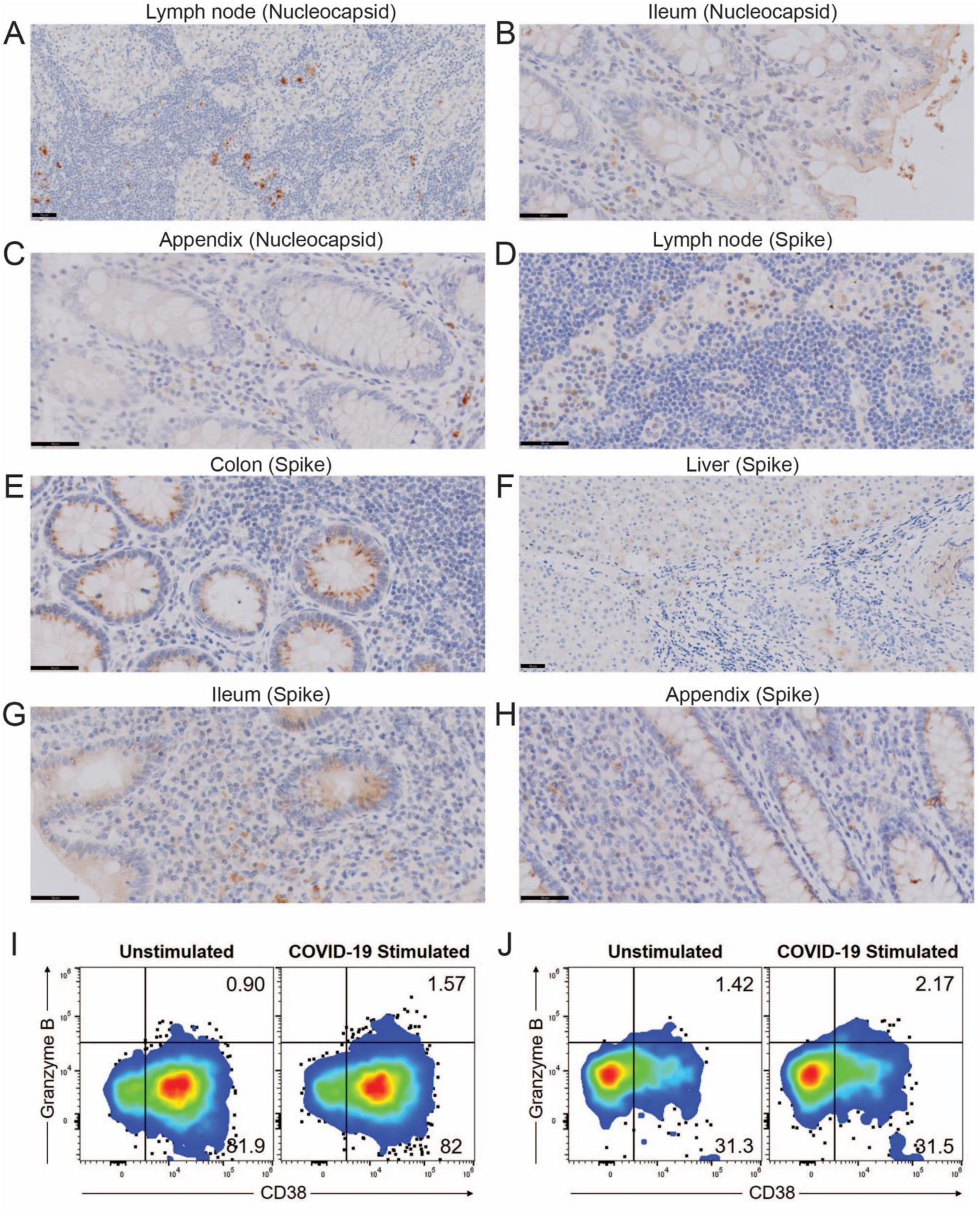
Immunohistochemistry and flow cytometry analysis of tissues obtained from two COVID-19 patients. (A) Positive SARS-CoV-2 nucleocapsid staining in scattered immune cells within the lymph node. (B-C) Positive SARS-CoV-2 nucleocapsid staining in the ileum (B) and the appendix (C). (D) Positive SARS-CoV-2 spike protein staining in scattered immune cells within the lymph node. (E-H) Positive SARS-CoV-2 spike protein staining in the colon (E), the liver (F), the ileum (G) and the appendix (H). Scale bar, 50 μm (A-H). (I-J) SARS-CoV-2-specific CD4^+^ T cells were identified from CD45^+^CD3^+^CD4^+^CD39^+^CD103^+^CD38^+^Granzyme B^+^ population, where CD39, CD103 and CD38 select for immune cells with memory phenotype (19, 20), and Granzyme B selects for immune cells with functional phenotype (21). Representative pseudocolour plots of CD38^+^Granzyme B^+^CD4^+^ T cells following stimulation with SARS-CoV-2 peptides in tissue (I) and blood (J) samples of patient 1. Similar results were obtained from patient 2. Numbers indicate percentages in the drawn gates. Plots shown are representative from one out of at least two independent experiments.

## Discussion

The SARS-CoV-2 viral antigen has been detected in the lung tissues of patients despite negative nasopharyngeal swab PCR tests (12). However, ours is the first study to detect the SARS-CoV-2 viral antigens in non-pulmonary tissues during the convalescent phase. Specifically, we detected residual SARS-CoV-2 in intestinal and liver tissues in two, clinically recovered COVID-19 patients. We have confidence in this finding, as we used the SARS-CoV-2 nucleocapsid antibody that was validated by the Centres for Disease Control and Prevention, which does not cross-react with other coronaviruses or common respiratory viruses (13). We also validated the results by RT-PCR and performed additional immunohistochemistry staining using antibody against the SARS-CoV-2 spike protein. Since the viral antigens were detected in these tissues, we interrogated whether the tissues show an immune response to the virus. Indeed, through *ex vivo* peptide stimulation, we observed SARS-CoV-2-specific T cell response from patients’ blood and tissues approximately 3 months into recovery; a finding of the tissues that we believe is the first to report to date.

The ACE2 receptor is necessary for viral entry, and it is highly expressed by intestinal epithelial cells. Thus unsurprisingly, we detected the viral antigens in ACE2-expressing cells in the colon. Incidentally, we also detected viral antigens in hepatocytes and sinusoidal Kupffer cells, suggesting that SAR-CoV-2 might indeed infect hepatocytes directly. Although many post-mortem studies have demonstrated SARS-CoV-2 in various organs (14), our findings from patients with mild COVID-19 are distinct from these post-mortem studies; patients who succumbed to COVID-19 had a more severe disease course and thus might not be representative of the general population.

Interestingly, several groups have reported the phenomenon where patients who had recovered from mild or moderate COVID-19 were later re-tested as positive in nasopharyngeal swabs or sputum samples (15, 16), raising concern for residual virus reservoirs and potential transmissibility in recovered individuals. Other studies have also reported positive SARS-CoV-2 RNA in anal swabs and stool samples, despite nasopharyngeal or sputum specimens testing negative for the virus (1, 3, 15, 17, 18). These reports are in line with our findings in colon tissues despite negative nasopharyngeal swab tests. Together, it seems that SARS-CoV-2 might be cleared in the digestive tract later than in the respiratory tract. Therefore, a negative nasopharyngeal swab result might not necessarily indicate complete viral clearance from the body.

Although respiratory transmission is responsible for most COVID-19 infections, faecal-oral transmission remains a possibility, with increasing evidence of COVID-19 causing gastrointestinal and hepatic manifestations, not to mention viral shedding in stools.^3^ Unfortunately, we were unable to determine whether the viral antigens isolated from the tissues were infectious, as the virus was inevitably destroyed during tissue fixation. We also did not have access to anal swab or stool samples, which would have provided additional insight into the possibility of faecal-oral transmission. Regardless, based on these preliminary findings, we believe that more research is warranted to understand the gastrointestinal and hepatic involvement in COVID-19.

Collectively, our findings constitute the first evidence for residual virus in extrapulmonary tissues during the convalescent phase, in a non-post-mortem setting. This phenomenon might have implications in terms of disease management and public health policy regarding transmission of COVID-19 infection besides respiratory route. We propose caution when handling tissues from patients who have a recent history of COVID-19, particularly during aerosol-generating procedures such as ultrasonic dissection surgery.

## Data Availability

All data are available from the corresponding authors upon reasonable request.

## Author Contributions

J.Y. and T.K.H.L. conceived, directed and supervised the study. T.T.T., S.K., J.L.N., and J.G.H.L. provided information and surgical samples of patient 1. P.C.C., C.Y.C., Y.X.K., and W.M.D.T. provided information and surgical samples of patient 2. T.Z.T. and J.C.T.L. performed histology-related technique. S.N.N. generated and provided figures. L.S. provided scientific inputs from pathology perspectives. C.C.L.C., D.G., and X.L. drafted the manuscript with the assistance of J.Y., with final reviews from all authors. All authors have read and agreed to the published version of the manuscript.

